# Acute Heart Failure in Pregnancy and Postpartum: Role of Iron Deficiency and Anemia in an Indian Multicenter Case–Control Study

**DOI:** 10.64898/2025.12.26.25343074

**Authors:** Manisha Nair, Saswati S Choudhury, Augustin Sundar, Sereesha Rao, Gitanjali Deka, Carolin Solomi, Anjali Rani, Vijay A Ismavel, Swapna D Kakoty, Pranabika Mahanta, Farzana Zahir, Indrani Roy, Rupanjali Deka, Charles Opondo, Megha Agarwal, Samuel Krasner, Paul Leeson, Samira Lakhal-Littleton, Barbara Casadei, Jane Armitage

**Affiliations:** Nuffield Department of Population Health, University of Oxford, UK & Department of Public Health, University of Copenhagen, Denmark; Department of Obstetrics and Gynecology, Gauhati Medical College and Hospital, Guwahati, Assam, India; Department of Obstetrics and Gynecology, Sewa Bhawan Hospital Society, Chattisgarh, India; Department of Obstetrics and Gynecology, Silchar Medical College and Hospital, Silchar, Assam, India; Department of Obstetrics and Gynecology, Makunda Christian Leprosy and General Hospital, Karimganj, Assam, India; Department of Obstetrics and Gynecology, Banaras Hindu University Institute of Medical Sciences, Varanasi, Uttar Pradesh, India; Department of Surgery, Makunda Christian Leprosy and General Hospital, Karimganj, Assam, India; Department of Community Medicine, Fakhruddin Ali Ahmed Medical College and Hospital, Barpeta, Assam, India; Department of Obstetrics and Gynecology, Jorhat Medical College and Hospital, Jorhat, Assam, India; Department of Obstetrics and Gynecology, Assam Medical College, Dibrugarh, Assam, India; Department of Obstetrics and Gynecology, Nazareth Hospital, Shillong, Meghalaya, India; MaatHRI Project, Srimanta Sankaradeva University of Health Sciences, Guwahati, Assam, India; Department of Medical Statistics, London School of Hygiene & Tropical Medicine, London, UK; Cardiovascular Clinical Research Facility, Radcliffe Department of Medicine, University of Oxford, UK; Department of Physiology, Anatomy and Genetics, University of Oxford, UK; National Heart and Lung Institute, Imperial College London, UK; Nuffield Department of Population Health, University of Oxford, UK

**Keywords:** Acute heart failure, pregnancy, postpartum, iron deficiency, anemia, peripartum cardiomyopathy, cardiovascular disease, India, low- and middle-income countries

## Abstract

**Background:** Cardiovascular diseases in pregnant and postpartum women remain under-researched, despite being the leading cause of indirect maternal deaths. In low- and middle-income countries (LMICs), anemia and iron deficiency are highly prevalent and may compound the cardiovascular stress of pregnancy, yet their contribution to maternal acute heart failure (AHF) is poorly understood. This study investigated the association between AHF in pregnancy/ postpartum and iron deficiency and anemia and assessed whether they remain consistent for peripartum cardiomyopathy (PPCM).

**Methods:** We conducted a multicenter case–control study in ten hospitals across four Indian states (February 2019–November 2024). Cases were pregnant/postpartum women with echocardiography-confirmed AHF as per standard definition; controls were postpartum women within 48 hours of childbirth without history/symptoms of heart disease recruited on the day of each case presentation. Blood samples collected at recruitment were analyzed for hemoglobin, ferritin, transferrin saturation (TSAT), soluble transferrin receptor (sTfR), and hepcidin. Multivariable logistic regression models assessed associations between iron status, anemia, and AHF, adjusting for potential confounders. Subgroup analyses were performed for PPCM.

**Findings:** Among 532 women with suspected heart failure, 317 had echocardiography-confirmed AHF and were compared with 1,091 controls. Moderate (Hb 7–9.9 g/dL) and severe (Hb <7 g/dL) anemia were respectively associated with 1.6-times (adjusted odds ratio [aOR] 1.59; 95% CI 1.10–2.32) and 6.1-times (aOR 6.09; 95% CI 3.35–11.07) the odds of AHF in women without anemia. Iron deficiency based on increasing levels of sTfR (aOR 1.47, 95% CI 1.32–1.63), lowest two hepcidin quintiles (aOR 4.07, 95% CI 2.26–7.32; aOR 2.06 95% CI 1.14–3.73) and low TSAT (aOR 1.50, 95% CI 1.01–2.21) were independently associated with AHF. Associations persisted after adjusting for hypertensive disorders of pregnancy and excluding women with pre-existing cardiac problems, and were consistent in the PPCM subgroup.

**Interpretation:** Iron deficiency and moderate-to-severe anemia substantially increase the risk of AHF in pregnancy and postpartum, independent of hypertensive or pre-existing conditions. These findings identify iron deficiency as a major, preventable, and under-recognized contributor to maternal cardiac complications in LMICs, highlighting the need for targeted antenatal screening and correction.

**CLINICAL PERSPECTIVE:** *What is new?:*

- This is the first multicenter study to demonstrate a strong, independent association between iron deficiency and acute heart failure during pregnancy and postpartum.
- The findings disentangle the effects of anemia and iron deficiency and confirm consistency across the PPCM subgroup.
- The findings identify iron deficiency with and without anemia as an important female-specific risk factor for pregnancy-associated acute heart failure.

*What are the clinical implications?:*

- Iron deficiency and anemia represent modifiable risk factors for maternal heart failure.
- Routine screening and timely correction of iron deficiency in pregnancy could reduce the risk of acute heart failure and related maternal deaths in LMICs.
- Integration of cardiac and iron parameter assessments should be prioritized in antenatal and postpartum care programs.

## Introduction

Cardiovascular diseases (CVD) in women remain under-researched, with evidence particularly limited for pregnant and postpartum women, despite CVD being the leading cause of indirect maternal deaths globally^1^. Notably, 92% of these deaths are considered preventable^2,3^. Pregnancy acts as a unique, sex-specific and time-limited physiological stressor that exposes underlying cardiovascular vulnerabilities. Consistent with this, reports suggest that the incidence of heart failure in pregnancy is rising^3^. During pregnancy, the maternal cardiovascular system undergoes profound adaptations including a 30–50% rise in cardiac output, systemic vasodilation, and plasma volume expansion that challenge vascular integrity and cardiac function^3^. These changes can unmask subclinical dysfunction (e.g., in hypertensive disorders of pregnancy (HDP) or gestational diabetes), exacerbate pre-existing cardiac disease (e.g., rheumatic or congenital heart disease), and magnify the adverse effects of physiological stressors such as iron deficiency.

Symptoms of heart failure in late pregnancy or the early postpartum period such as fatigue, breathlessness, and oedema are often overlooked as normal physiological changes. Heart failure is a syndrome. It can be caused by reduced left ventricular ejection fraction (LVEF) but also by the hyperdynamic circulation and relative hypoxia caused by anemia. In other words, severely anemic individuals could have heart failure, even though they do not necessarily have reduced LVEF. Anemia affects more than a third of all pregnancies worldwide^4^ and is associated with increased maternal mortality, particularly in low- and middle-income countries (LMICs)^5^. Pregnant women experience substantial cardiovascular stress^6^, revealing latent predispositions to CVD in anemic women through the physiological demands it imposes^7^. The European Society of Cardiology (ESC) identifies anemia as a precipitating factor for acute heart failure (AHF)^8^. Despite this, there is limited evidence about cardiovascular risks associated with anemia in pregnancy particularly in LMIC settings^9^.

In an Indian population, we estimated the incidence of AHF in pregnancy and postpartum to be ∼2 per 1,000 hospital births, with a case-fatality of 40%^10^. Clinical observations raised suspicion that iron deficiency anemia contributed to this high risk in a setting where iron deficiency anemia is highly prevalent, affecting 34-55% women during pregnancy and postpartum with hemoglobin levels as low as 2 g/dL^11^. The pathophysiology of AHF is heterogeneous, with multiple precipitating factors capable of triggering new-onset or acute worsening of pre-existing cardiac disease^12^. Our systematic review found no human studies directly investigating the role of iron deficiency in AHF during pregnancy or postpartum. However, a meta-analysis of five studies showed that women with peripartum cardiomyopathy (PPCM) had lower hemoglobin levels compared with controls^13^. Additionally, three published papers from India reported a 27–52% prevalence of heart failure in women with severe anemia (hemoglobin <7gm/dL)^14–16^. Here we investigated the association between AHF in pregnancy/ postpartum and maternal iron deficiency and anemia and assessed whether these associations remain consistent in women with PPCM.

## Methods

### Study design and population

We conducted an unmatched case–control study from February 2019 to November 2024 in ten hospitals across four Indian states through the Maternal and Perinatal Health Research Collaboration, India (MaatHRI)^17^.

Cases were pregnant or postpartum women presenting with AHF at any time during pregnancy, childbirth, or up to 6 months postpartum. AHF is defined as “the new onset (de novo heart failure) or worsening (acutely decompensated heart failure) of symptoms and signs of HF, mostly related to systemic congestion”^8,12^. Controls were postpartum women within 48 hours of childbirth, without history or symptoms of cardiac disease, recruited on the same day of a case presenting at the hospital.

To improve case identification, a broad screening criteria was used based on the known signs and symptoms of AHF^12^ and adapting the California Quality Improvement Toolkit on cardiovascular screening and assessment for pregnant women^18^. Women were diagnosed with AHF if they presented de novo with breathlessness (respiratory rate ≥15/min) and one or more of the following symptoms: elevated jugular venous pressure, cardiac murmur, or pulmonary oedema. AHF was then confirmed by evidence of cardiac dysfunction on transthoracic echocardiography, performed using a validated focused cardiac ultrasound protocol described previously^19^. Echocardiography was performed in all cases and all controls recruited from October 2019 onwards. Due to an initial shortage of trained sonographers, echocardiography of control participants started at a later time-point than the cases.

A subset of the confirmed cases was classified as PPCM, defined as “an idiopathic cardiomyopathy presenting with heart failure secondary to left ventricular systolic dysfunction (LVEF <45%) towards the end of pregnancy or in the months following delivery, in the absence of other causes”^20,21^. Brain Natriuretic Peptide (BNP) was not measured as values are influenced by pregnancy and hypertensive disorders of pregnancy (HDP) which had a prevalence of 8.3% in our population^3^; the test was therefore considered cost-ineffective in this setting.

### Inclusion and exclusion criteria

Incident cases that had a confirmed abnormality detected by echocardiography were included as confirmed AHF cases. Suspected cases without echocardiographic evaluation were excluded. For PPCM subgroup analyses, cases not fulfilling PPCM diagnostic criteria were excluded. Women unwilling or unable to provide informed consent (or for whom consent from next of kin was not available) were excluded.

### Baseline and follow-up data

In addition to echocardiography, non-fasted blood samples were collected at recruitment from cases at hospital admission and from controls within 48 hours of childbirth. Laboratory methods are described in the supplementary file. Standard data collection forms were used to collect baseline information about socio-demographic (maternal age, body mass index (BMI), religion, residence, education level, and living below the poverty line status), behavioral/ lifestyle (smoking status, alcohol consumption, tobacco consumption and chewing betelnut), obstetric factors (parity, multiple pregnancy, HDP, antenatal check-ups), and pre-existing medical comorbidities. These are described in detail in our previous papers^22,23^. We also collected information about hemoglobin measures from the antenatal records using the earliest available value; other iron parameters were unavailable in antenatal records. Information about blood transfusion prior to AHF onset was recorded.

Additional clinical information from the cases included blood pressure, New York Heart Association Functional Classification (NYHA) based on severity of fatigue, palpitation and dyspnea for grading severity of the heart failure symptoms^24^, tachycardia, timing of HF onset, and history of cardiac problems. All clinical and obstetric data were collected from hospital records and cases and controls were followed up to record information about labor, childbirth and fetal outcomes. Women were again contacted at 6 weeks postpartum for outcomes including rehospitalization, complications, and mortality. Written informed consent was obtained; if the woman was too unwell, next-of-kin consent was accepted.

### Anemia and iron status (exposure variables)

Anemia was defined using the World Health Organization (WHO) definition for hemoglobin cut-offs: mild anemia, hemoglobin 10–10.9 g/dL; moderate 7–9.9 g/dL; and severe <7 g/dL^25^.

Similar to our previous study^11^, iron deficiency was defined using standard biomarkers. sTfR, an indicator of iron availability to the bone marrow for erythropoiesis, was used as a continuous variable as there is no agreed physiological cut-off, with higher levels suggestive of unmet iron need. sTfR is not known to be influenced by inflammation or pregnancy. TSAT, a measure of circulating serum iron available to tissues, was classified as <16%, 16%–50%, and >50% to respectively denote low, normal, and high levels relevant to our study population as described in our previous study^11^. Serum hepcidin is a hormone that both regulates and is regulated by iron availability in the body. It was categorized into quintiles as there is no recommended physiological cut-off for hepcidin in pregnancy.

Serum ferritin is a measure of body iron stores but it is also a positive acute phase protein increased by inflammation due to which there is a lack of consensus on the cut-offs for low and high ferritin levels in patients with heart failure^26^. Therefore, we used ferritin as a secondary biomarker instead of excluding it from our study as it is commonly used as a measure of iron status in LMICs. The supplementary materials include a description of the methodology for adjusting serum ferritin values for inflammation^27^. To avoid misclassification of iron deficient and iron overload individuals, WHO recommends defining low and high ferritin levels in populations within the context of the extent of inflammation and infection^28^. The ESC defines iron deficiency in HF patients taking into account the extent of inflammation as serum ferritin concentration <100 ng/mL, or 100–299 ng/mL with TSAT <20%^29^. Therefore, low ferritin in the AHF cases were defined using the ESC guidance and low ferritin in control population was defined using a standard approach of <30 ng/mL^11,30,31^. Ferritin values above 95^th^ percentile were categorized as high ferritin for both cases and controls. This was considered to be the most appropriate approach due to the known challenges related to cut-offs for ferritin in pregnancy and postpartum in populations more likely to be exposed to infections and inflammation and in this study also presenting with AHF.

### Study power

Assuming a 25% prevalence of iron deficiency anemia^11^, a 3:1 control-to-case ratio, 80% power, 5% alpha, and odds ratio of 1.5, 250 confirmed cases were required. Allowing 20% attrition, the target sample size was 300 cases and 900 controls. To achieve this, we planned to screen ∼500 suspected AHF cases.

### Statistical analysis

Socio-demographic, behavioral and obstetric characteristics of the confirmed cases and controls were compared using descriptive statistics. Echocardiography findings of all suspected cases and controls, and clinical characteristics of the confirmed AHF cases were described. Continuous variables were summarized as medians (IQR), categorical variables as proportions. Non-linear associations were observed between hemoglobin (at recruitment and earliest antenatal visit) and AHF (supplementary Figures S3 and S4); hemoglobin was therefore categorized by anemia severity.

Multivariable logistic regression was used to examine independent associations between blood parameters and AHF, adjusting for confounders, clustering by hospital and gestational age/postpartum days at blood draw. Regular, rather than conditional, logistic regression was used as this was an unmatched case-control study. The multivariable model for TSAT was additionally adjusted for blood transfusion in the antenatal period prior to the AHF onset, and ferritin and hepcidin models were additionally adjusted for CRP to control for inflammation. We re-ran the analysis by excluding 30 women with a known history of cardiac problem. Interaction terms and likelihood ratio tests were used to investigate interaction between iron parameters/ anemia and HDP or iron parameters/ anemia and pre-medical comorbidities in their effect on AHF. If evidence of interaction was found, a stratified analysis was conducted. We also explored the independent effect of the iron parameters stratified by anemia (no/mild and moderate-severe anemia). These analyses were repeated for the sub-group analysis comparing PPCM cases and controls.

Data was assumed to be missing at random and complete case analysis was performed. However, blood collection was not feasible in some severely unwell AHF cases including in 28 of 30 women who died, thus missingness for the blood parameters could be related to severity of the outcome. We therefore conducted a sensitivity analysis categorizing women with missing data as a separate sub-group. Analyses were conducted using Stata v18.5 MP (StataCorp).

### Data availability statement

The data collected for this study are sensitive data and are therefore available from the corresponding author upon reasonable request.

### Ethics approvals

This MaatHRI study was approved by the institutional review boards of each participating Indian coordinating institution, namely: Srimanta Sankaradeva University of Health Sciences, Guwahati, Assam (No.MC/190/2007/Pt-1/126); Nazareth Hospital, Shillong, Meghalaya (Ref No. NH/CMO/IEC/COMMUNICATIONS/18-01); Emmanuel Hospital Association, New Delhi (Ref. Protocol No.167); and the Institute of Medical Sciences, Banaras Hindu University, Varanasi, Uttar Pradesh (No.Dean/2018/EC/290). The studies were approved by the Government of India’s Health Ministry’s Screening Committee, the Indian Council of Medical Research, New Delhi (ID number 2018-0152) and by Oxford Tropical Research Ethics Committee, University of Oxford, UK (OxTREC Ref: 7-18).

## Results

Of 532 women with suspected AHF based on clinical screening, 425 received echocardiographic assessment. The remaining 107 were excluded: 10 due to lack of consent, 3 died before assessment, and 94 did not receive echocardiography due to unavailability of trained clinicians or technical issues with image sharing. Median levels of hemoglobin and iron parameters did not differ significantly between suspected cases with and without echocardiographic evaluation. Among the 425 assessed women, 317 (75%) had cardiac dysfunction detected by echocardiography and were confirmed as AHF. Of these, 66 met the diagnostic criteria for PPCM. A data flow diagram for the cases is presented in Figure-1.

A total of 1,091 controls were enrolled, of whom 644 underwent echocardiographic assessment, mainly due to the delayed start of echocardiography in controls as explained above but some due to technical issues with image sharing. All 1,091 were included in the analysis. Follow-up at 6 weeks postpartum was achieved for 90% of cases and 94% of controls.

### Characteristics of the study population

The characteristics of the confirmed cases (AHF and PPCM) and controls are shown in Table-1. Compared to the controls, a higher proportion of women who had AHF were 35 years or older, had lower BMI (<18.5 kg/m^2^), belonged to Muslim or other minority religious backgrounds, lived in rural areas, and were illiterate or had only primary education. Socioeconomic status measured by women’s below poverty line status did not vary between cases and controls. In terms of behavioral characteristics, very few women smoked or consumed alcohol, but a higher proportion of cases than controls consumed tobacco or chewed betelnut during pregnancy or gave up just before pregnancy. A significantly higher proportion of cases were multiparous, had twin pregnancy and received three or fewer antenatal check-ups. Compared with controls, a higher proportion of women with AHF were diagnosed with HDP and had pre-existing medical problems including primarily history of cardiac problems (n=30, see Table-2), but also asthma (n=2), hypothyroidism (n=1) and pneumonia (n=2).

**Table 1:** Socio-demographic, behavioural and obstetric characteristics of the study population.

| Characteristics of study participants<sup>‡</sup> | Controls (n=1091) | AHF (n=317) | p-value* | PPCM case (n=66) | p-value** |
| --- | --- | --- | --- | --- | --- |
| **Socio-demographic** |  |  |  |  |  |
| Age (in years) |  |  |  |  |  |
| <20 | 135 (12.4) | 38 (12.0) | <0.001 | 12 (18.2) | <0.001 |
| 20-24 | 405 (37.1) | 110 (34.7) |  | 19 (28.8) |  |
| 25-29 | 373 (34.2) | 87 (27.4) |  | 14 (21.2) |  |
| 30-34 | 142 (13.0) | 47 (14.8) |  | 12 (18.2) |  |
| ≥35 | 36 (3.3) | 33 (10.4) |  | 8 (12.1) |  |
| BMI in (kg/m<sup>2</sup>) |  |  |  |  |  |
| Underweight (BMI <18.5) | 68 (6.2) | 55 (17.3) | <0.001 | 12 (18.2) | <0.001 |
| Normal (BMI 18.5 to 24.9) | 813 (74.5) | 198 (62.5) |  | 42 (63.6) |  |
| Overweight/ obese (BMI ≥25) | 161 (14.8) | 37 (11.7) |  | 6 (9.1) |  |
| Religion |  |  |  |  |  |
| Hindu | 765 (70.1) | 179 (56.5) | <0.001 | 29 (43.9) | <0.001 |
| Muslim | 298 (27.3) | 122 (38.5) |  | 30 (45.5) |  |
| Others | 28 (2.6) | 14 (4.4) |  | 6 (9.1) |  |
| Residence |  |  |  |  |  |
| Rural | 767 (70.3) | 253 (79.8) | <0.001 | 61 (92.4) | <0.001 |
| Urban/ semi-urban | 324 (29.7) | 62 (19.6) |  | 4 (6.1) |  |
| Living below the poverty line |  |  |  |  |  |
| No | 323 (29.6) | 95 (30.0) | 0.322 | 14 (21.2) | 0.082 |
| Yes | 684 (62.7) | 196 (61.8) |  | 50 (75.8) |  |
| Not known | 84 (7.7) | 25 (7.9) |  | 2 (3.0) |  |
| Level of education |  |  |  |  |  |
| Illiterate | 47 (4.3) | 38 (12.0) | <0.001 | 15 (22.7) | <0.001 |
| Primary | 114 (10.4) | 49 (15.4) |  | 14 (21.2) |  |
| Secondary | 744 (68.2) | 188 (59.3) |  | 34 (51.5) |  |
| Higher | 185 (17.0) | 39 (12.3) |  | 2 (3.0) |  |
| **Lifestyle/ behavioural factors** |  |  |  |  |  |
| Smoking status |  |  |  |  |  |
| Never smoked | 1087 (99.6) | 312 (98.4) | 0.010 | 65 (98.5) | 0.001 |
| Smoked prior to/ during pregnancy | 4 (0.4) | 2 (0.6) |  | 0 |  |
| Alcohol consumption |  |  |  |  |  |
| Never | 1084 (99.4) | 311 (98.1) | 0.013 | 65 (98.5) | 0.001 |
| Consumed prior to/ during pregnancy | 7 (0.6) | 3 (1.0) |  | 0 |  |
| Tobacco consumption |  |  |  |  |  |
| Never/ prior to pregnancy | 1048 (96.1) | 290 (91.5) | <0.001 | 54 (81.8) | <0.001 |
| Current/ gave up during pregnancy | 43 (3.9) | 24 (7.6) |  | 11 (16.7) |  |
| Chewing betelnut |  |  |  |  |  |
| Never/ prior to pregnancy | 599 (54.9) | 151 (47.6) | 0.001 | 24 (36.4) | <0.001 |
| **Obstetric and medical history** |  |  |  |  |  |
| Parity |  |  |  |  |  |
| Primiparous | 747 (68.5) | 189 (59.6) | <0.001 | 41 (62.1) | <0.001 |
| Multiparous (1-2 previous pregnancies) | 316 (28.9) | 103 (32.5) |  | 16 (24.2) |  |
| Multiparous (≥3 previous pregnancies) | 28 (2.6) | 23 (7.3) |  | 8 (12.1) |  |
| Received antenatal check-ups |  |  |  |  |  |
| None | 16 (1.5) | 15 (4.7) | <0.001 | 5 (7.6) | <0.001 |
| ≤ 3 check-ups | 361 (33.1) | 174 (54.9) |  | 44 (66.7) |  |
| > 3 check-ups | 714 (65.4) | 124 (39.1) |  | 16 (24.2) |  |
| Multiple (twin) pregnancy |  |  |  |  |  |
| No | 1077 (98.7) | 301 (94.9) | <0.001 | 63 (95.5) | <0.001 |
| Yes | 14 (1.3) | 13 (4.1) |  | 2 (3.0) |  |
| Hypertensive disorders of pregnancy |  |  |  |  |  |
| No | 962 (88.2) | 198 (62.5) | <0.001 | 35 (53.0) | <0.001 |
| Yes | 129 (11.8) | 117 (36.9) |  | 30 (45.5) |  |
| Any pre-existing medical problems |  |  |  |  |  |
| No | 1052 (96.4) | 273 (86.1) | <0.001 | 62 (93.9) | 0.469 |
| Yes | 24 (2.2) | 35 (11.0) |  | 3 (4.6) |  |

\*p-value for Chi<sup>2</sup> test for difference in proportion comparing AHF and controls; \*\*p-value for Chi<sup>2</sup> test for difference in proportion comparing PPCM sub-group and controls; ‡ less than 3% missing information except for BMI: 4.5% (n=49) missing for controls and 8.5% (n=27) missing for cases.

**Table 2:** Clinical characteristics of women with acute heart failure (AHF) and peripartum cardiomyopathy (PPCM)

| Characteristic | AHF cases (n = 317) | PPCM cases (n = 66) |
| --- | --- | --- |
| **Blood pressure\*, mean (range), mmHg** |  |  |
| Systolic | 130.6 (60–220) | 135.5 (80–200) |
| Diastolic | 84.7 (50–170) | 87.8 (50–120) |
| **NYHA classification, n (%)** |  |  |
| Grade I (no limitation) | 21 (6.6) | 1 (1.5) |
| Grade II (slight limitation) | 65 (20.5) | 10 (15.1) |
| Grade III (marked limitation) | 39 (12.3) | 9 (13.6) |
| Grade IV (symptoms at rest) | 189 (59.6) | 46 (69.7) |
| Not known | 3 (1.0) | 0 (0) |
| **Presenting with tachycardia, n (%)** |  |  |
| No | 237 (74.8) | 48 (72.7) |
| Yes | 53 (16.7) | 16 (24.2) |
| Not known | 27 (8.5) | 2 (3.0) |
| **Onset of HF, n (%)** |  |  |
| Antenatal | 159 (50.2) | 27 (40.9) |
| Median gestational age, weeks (IQR) | 34 (30–37) | 36 (30–39) |
| Labour/delivery | 25 (7.9) | 3 (4.6) |
| Postpartum | 130 (41.0) | 36 (54.5) |
| Median onset postpartum (IQR) | 48 h (15 h–8 d) | 6.8 d (1.3–14.5) |
| Not known | 3 (0.9) | 0 (0) |
| **Outcome, n (%)** |  |  |
| Survived | 235 (74.1) | 48 (72.7) |
| Died<sup>‡</sup> | 25 (7.9) | 12 (18.2) |
| Not known | 57 (18.0) | 6 (9.1) |
| **History of cardiac problems, n (%)** |  |  |
| None | 284 (89.6) | *No known history, by PPCM definition* |
| Rheumatic heart disease | 26 (8.2) |  |
| Congenital heart disease | 2 (0.6) |  |
| Other (not specified) | 2 (0.6) |  |
| Not known | 3 (1.0) |  |

\*Blood pressure recordings not available for 5 AHF cases; NYHA = New York Heart Association; “Not known” reflects missing data; <sup>‡</sup>Died’ group includes those with confirmed AHF on echocardiographic assessment.

### Echocardiographic findings of suspected cases and controls

The most common echocardiographic abnormality among women with AHF was pericardial effusion, present in one-third of cases, of which more than 80% were of moderate-to-severe grade (supplementary Table S1). The pericardial fluid appeared clear in all echocardiograms except for one case. Valvular involvement was also frequent: mitral regurgitation was detected in 31% (19% mild and 12% moderate-to-severe), while tricuspid regurgitation was observed in 22% (14% mild and 8% moderate-to-severe). Chamber enlargement was identified as 19% for the left atrium, 12% for the left ventricle, and around 10% each for the right atrium and right ventricle. Left ventricular systolic function was preserved in the majority, but approximately 30% of cases had a reduced LVEF (<55%) and in 11% of cases left ventricular regional wall motion abnormalities were detected.

Among the 644 controls who underwent echocardiographic evaluation, 11.7% (75 of the 644 controls) had significant findings, the most common being pericardial effusion (supplementary Table S1). However, unlike the cases, these women were asymptomatic with no history of cardiac problems and had not been suspected of heart failure at the time of recruitment.

### Clinical presentation of confirmed heart failure cases

Baseline clinical characteristics of cases are shown in Table-2. Half of the AHF cases presented antenatally (median gestational age 34 weeks), 40% in the early postpartum period (median 48 hours after delivery), and 8% during labor or childbirth. Most cases (72%) were classified as NYHA Grade III or IV, indicating severe functional limitation. 30 women presenting with suspected HF died after recruitment (25 confirmed AHF, 2 unconfirmed and 3 with no echocardiography assessment). Notably, 90% of cases had no known history of cardiac disease. A history of rheumatic heart disease was reported by 8%, whereas echocardiography identified rheumatic valve characteristics in 13% (supplementary Table S1).

PPCM cases showed broadly similar clinical features (Table-2). However, a greater proportion presented in the postpartum period, with a median onset approximately one week after delivery. The majority (83%) were classified as NYHA Grade III or IV.

### Description of iron parameters in the study population

Compared with controls, median hemoglobin levels were lower in women who had AHF with 17% and 18% having severe anemia at recruitment and at earliest antenatal visit, compared with 4% and 2% among controls, respectively (Table-3). The prevalence of abnormal hemoglobin types (HbE trait and homozygous, β-thalassemia trait, sickle cell trait or unspecified) was ∼20% in both groups, although a higher proportion of women with AHF (11.4%) did not have information about their hemoglobin variant compared with 3% of the controls. The iron parameters indicated a higher prevalence of iron deficiency among cases than controls. Median sTfR levels were significantly higher and median hepcidin levels were lower among cases (26% of cases were in the lowest hepcidin quintile compared with 15% of controls). However, median TSAT levels were comparable between cases and controls although a higher proportion of AHF cases had high TSAT >50% at recruitment. Secondary analysis of serum ferritin showed higher proportion of low ferritin among cases than controls if we use the ESC definition, but if we use the inflammation corrected values alone, the proportion of women with low ferritin appears to be lower and high ferritin appears to be higher among cases compared with controls. This suggests that the effect of inflammation in AHF cases could not be accounted for by the adjustments alone. The comparison of the levels of blood parameters and the proportions of anemia and iron deficiency in the PPCM cases and controls was very similar (Table-3).

**Table 3:**
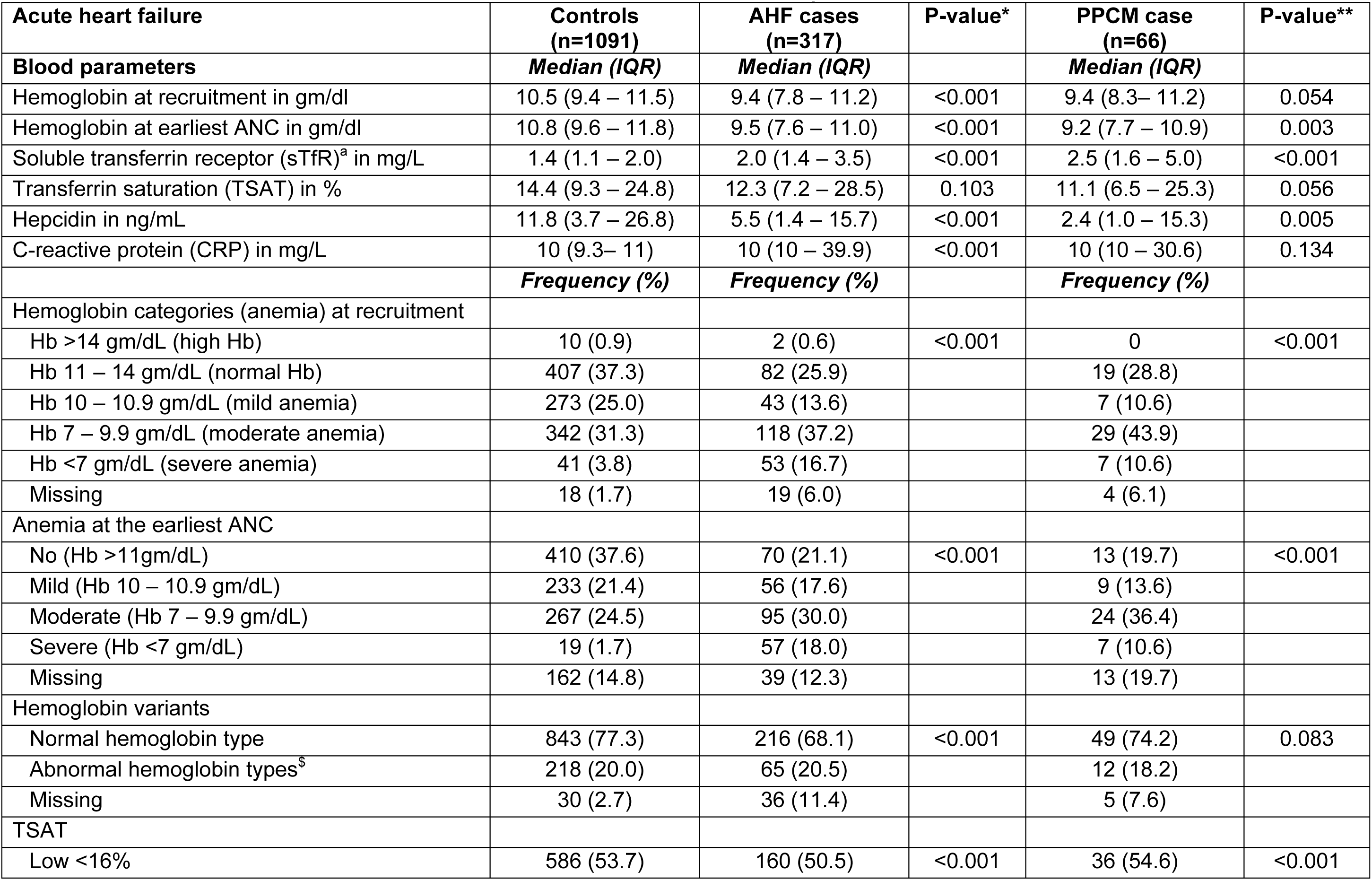
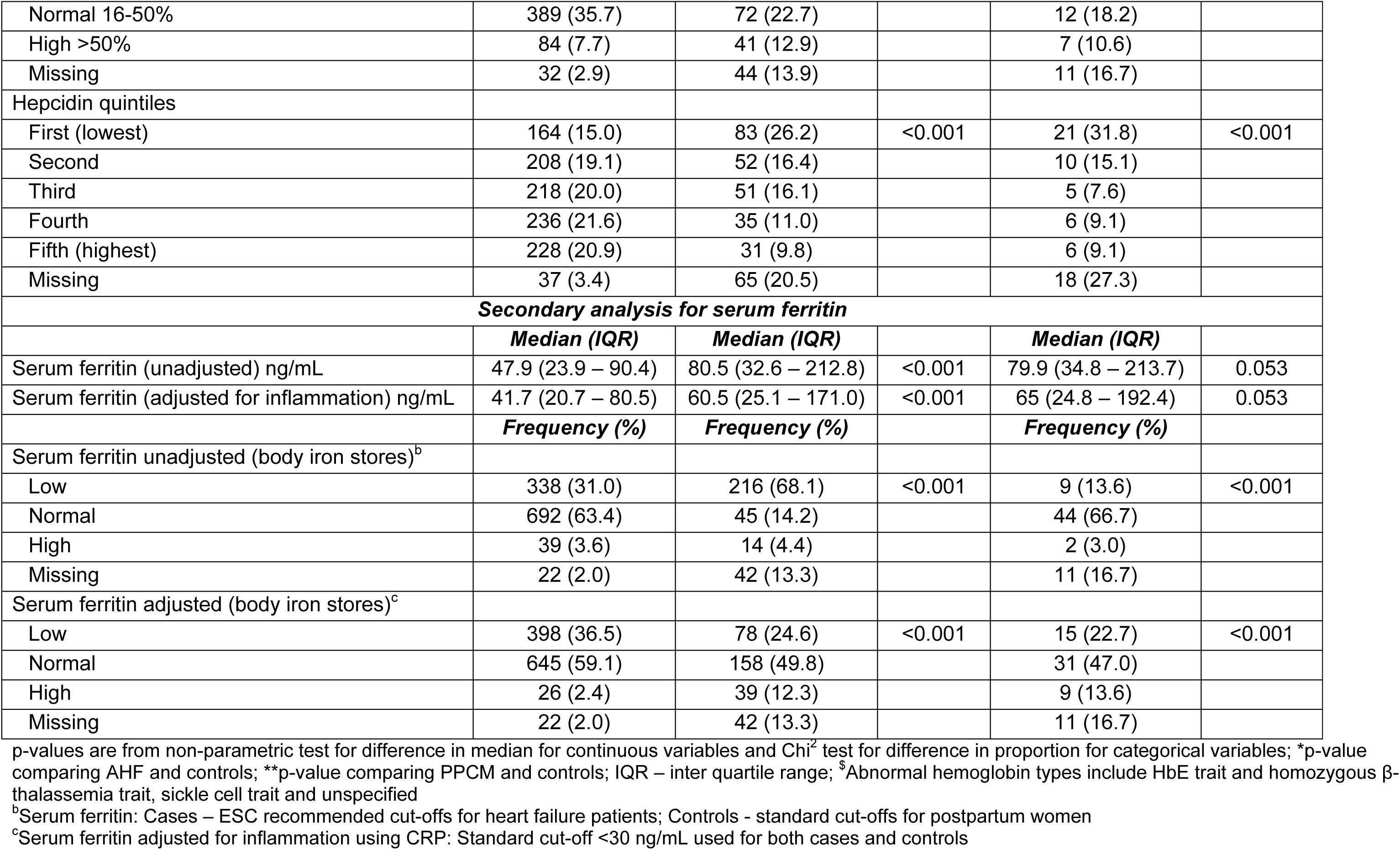
Biomarkers and clinical characteristics of AHF and PPCM cases compared with controls.

Among the controls who had a significant cardiac dysfunction on echocardiographic evaluation 41.3% (31 of 75 women) had moderate-severe anemia, 36% (n=27) had low serum ferritin, 57.3% (n=43) had low TSAT, 36% (n=27) were in the lowest two hepcidin quintiles, 12% (n=9) had HDP and one woman had a non-cardiac medical comorbidity.

Furthermore, comparing the echocardiographic findings between women who had iron deficiency and those who did not, showed a higher proportion of valvular defects and both left and right ventricular enlargements in the iron-deficient groups compared with the non-deficient group, but the proportions were not statistically significantly (Table S2).

### Effect of maternal anemia and hemoglobin variants on AHF

After adjusting for potential socio-demographic, behavioral, medical and obstetric confounders, clustering by hospitals and days from conception to blood collection, we found 60% higher odds of AHF among women who had moderate anemia (adjusted odds ratio (aOR) 1.59, 95% CI 1.10 - 2.32, p=0.014) and more than 6 times the odds of AHF among women with severe anemia (aOR 6.09, 95% CI 3.35 - 11.07, p<0.001) compared with women who did not have anemia at recruitment (Figure 2 and supplementary Table S3). The odds of AHF did not vary significantly between women with mild and no anemia. The effect estimates were even larger for the association between AHF and moderate and severe anemia at the earliest antenatal visit prior to AHF onset (Figure 2 and Table S3). We did not find a significant association between AHF and hemoglobin variants (aOR 1.13, 95% CI 0.73 - 1.74, p=0.585; Figure 2 and Table S3). Re-running the analyses by excluding 30 women with a known history of cardiac problem did not change the findings (*results not shown*).

**Figure 1:**
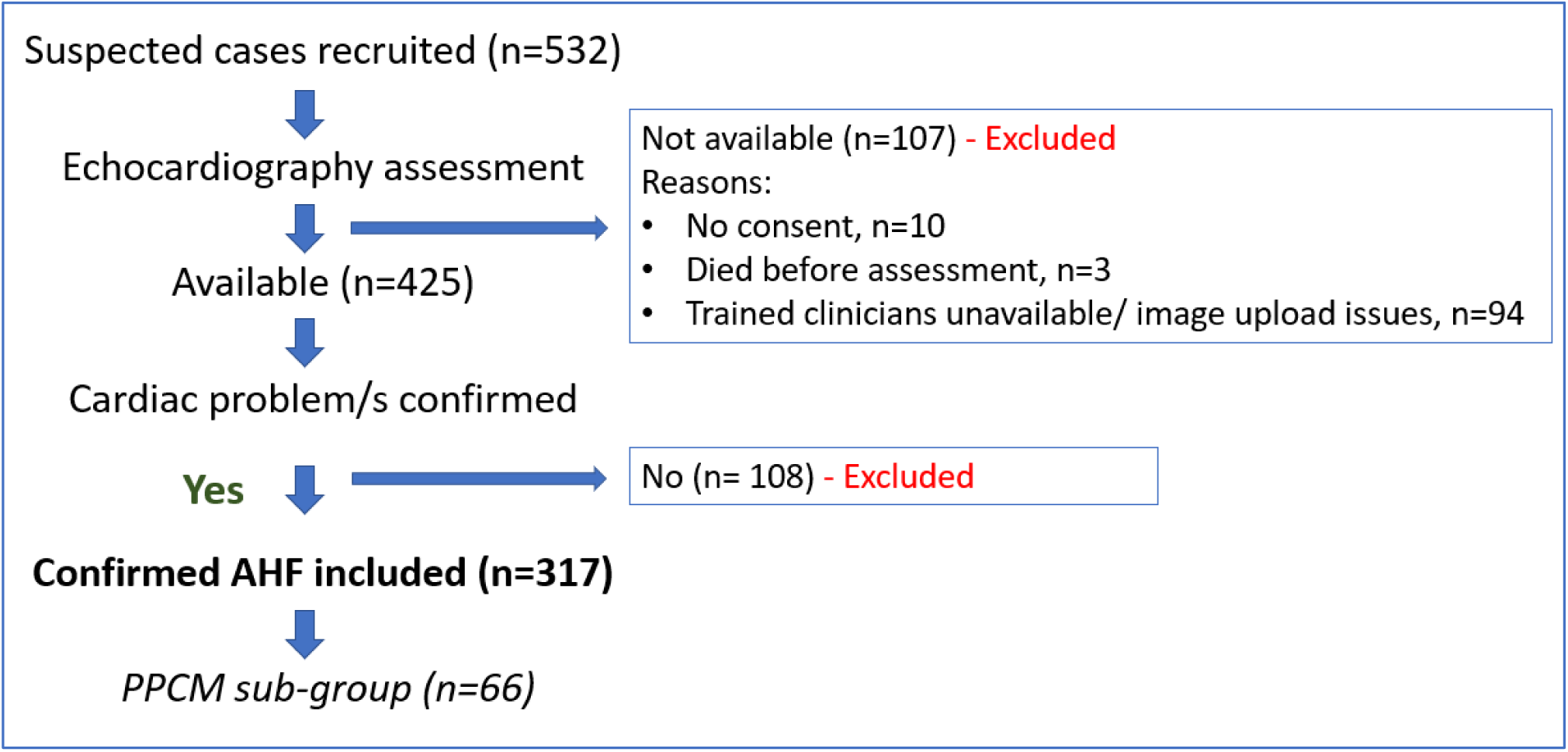
Data flow diagram for acute heart failure cases

**Figure 2:**
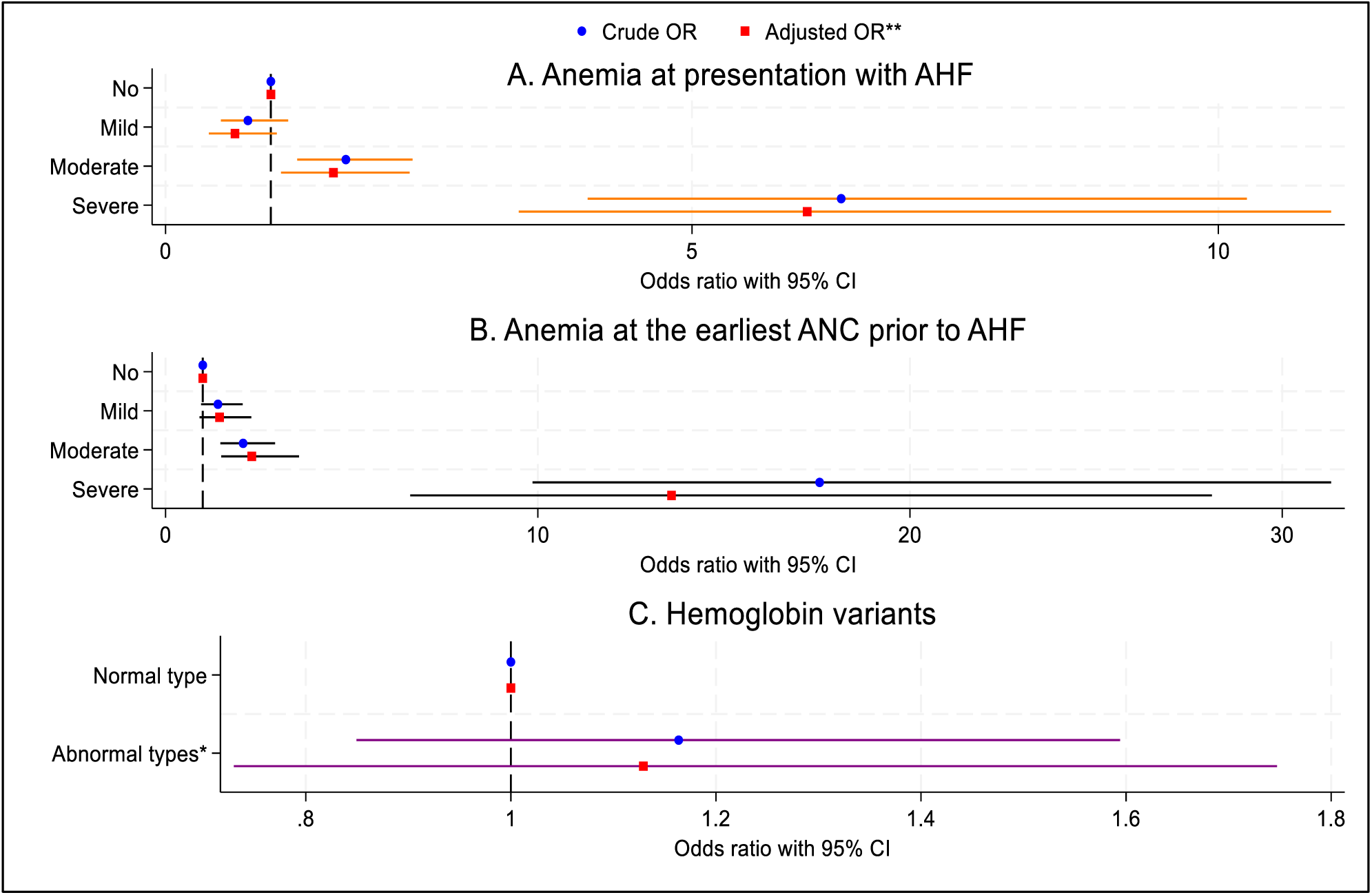
Association of maternal anemia and hemoglobin variants with AHF Notes: different scales for the OR in the three figures. Anemia severity for figures A and B is defined as: No anemia Hb >11gm/dL, Mild anemia Hb 10 – 10.9 gm/dL; Moderate anemia Hb 7 – 9.9 gm/dL; and Severe anemia Hb <7 gm/dL. **Adjusted for woman’s age, BMI, religion, residence, below poverty line status, education level, tobacco, alcohol and betelnut consumption, smoking status, parity, antenatal care visits, medical comorbidities, multiple pregnancy, clustering by hospitals, days from conception to blood collection (all three figures). *Abnormal hemoglobin types (Figure C) include HbE trait and homozygous β-thalassemia trait, sickle cell trait and unspecified.

There was evidence of an interaction between anemia and HDP on their effect on AHF (interaction p=0.0209) but no evidence of interaction between anemia and medical comorbidities (p=0.1334) after controlling other factors. Therefore, HDP was not adjusted for as a confounder, instead a stratified analysis by presence or absence of HDP was conducted in addition to the primary analysis. There was no evidence of interaction between either factor and anemia at the earliest antenatal visit (HDP and antenatal anemia interaction p=0.0360; medical comorbidities and antenatal anemia interaction p=0.2787). Additionally, there was no evidence of interaction between hemoglobin variants and HDP (interaction p=0.1681) or hemoglobin variants and medical comorbidities (interaction p=0.0501) in their effects on AHF, therefore HDP and medical comorbidities were adjusted for as confounders in the analysis investigating the association between AHF and hemoglobin variants.

The stratified analysis by presence or absence of HDP further showed evidence of adjusted increased odds of AHF among pregnant and postpartum women with moderate and severe anemia among women who did not have HDP (supplementary Table S4) with larger effect estimates. Importantly, women who had mild anemia at the earliest antenatal visit before AHF also had a significantly higher odds of AHF (compared with women with no anemia) if they did not have HDP (Table S4).

The association between PPCM and anemia and hemoglobin variants were similar to that of AHF (see Table S5). Unlike AHF, no evidence of interaction was observed in the PPCM analysis.

### Effect of iron parameters on AHF

After adjusting for socio-demographic, behavioral, obstetric and medical factors, clustering by hospitals and timing of blood collection in relation to pregnancy, we found a positive linear association between sTfR and AHF (Figure 3) with odds of AHF increasing by 47% per unit increase in sTfR (95% CI 32% - 63%, p<0.001; Figure-4.A and supplementary Table S6). Increasing levels of sTfR are suggestive of unmet iron need and absolute iron deficiency. No non-linear associations between AHF and sTfR were observed. Low TSAT, indicating iron deficiency, was associated with 50% higher odds of AHF after controlling for potential confounders including blood transfusion (aOR 1.50, 95% CI 1.01 - 2.21, p=0.045; Figure 4.B and Table S6). The unadjusted higher odds of AHF observed among women with high TSAT (>50%) was attenuated and there was no evidence of association after adjusting for confounders, specifically blood transfusion in the antenatal period. The odds of AHF in the lower two (first and second) quintiles of hepcidin were two- and four-times the odds of AHF in the highest (fifth) quintile after adjusting for confounders including inflammation, but the second highest and the highest quintiles were not different (Fourth compared with fifth quintile aOR 1.23, 95% CI 0.66 – 2.30, p=0.517; third vs fifth quintile aOR 1.77, 95% CI 0.98 – 3.20, p=0.060; second vs fifth aOR 2.06, 95% CI 1.14 – 3.73, p=0.017; first vs fifth aOR 4.07, 95% CI 2.26 – 7.32, p<0.001; Figure 4.C and Table S6).

**Figure 3:**
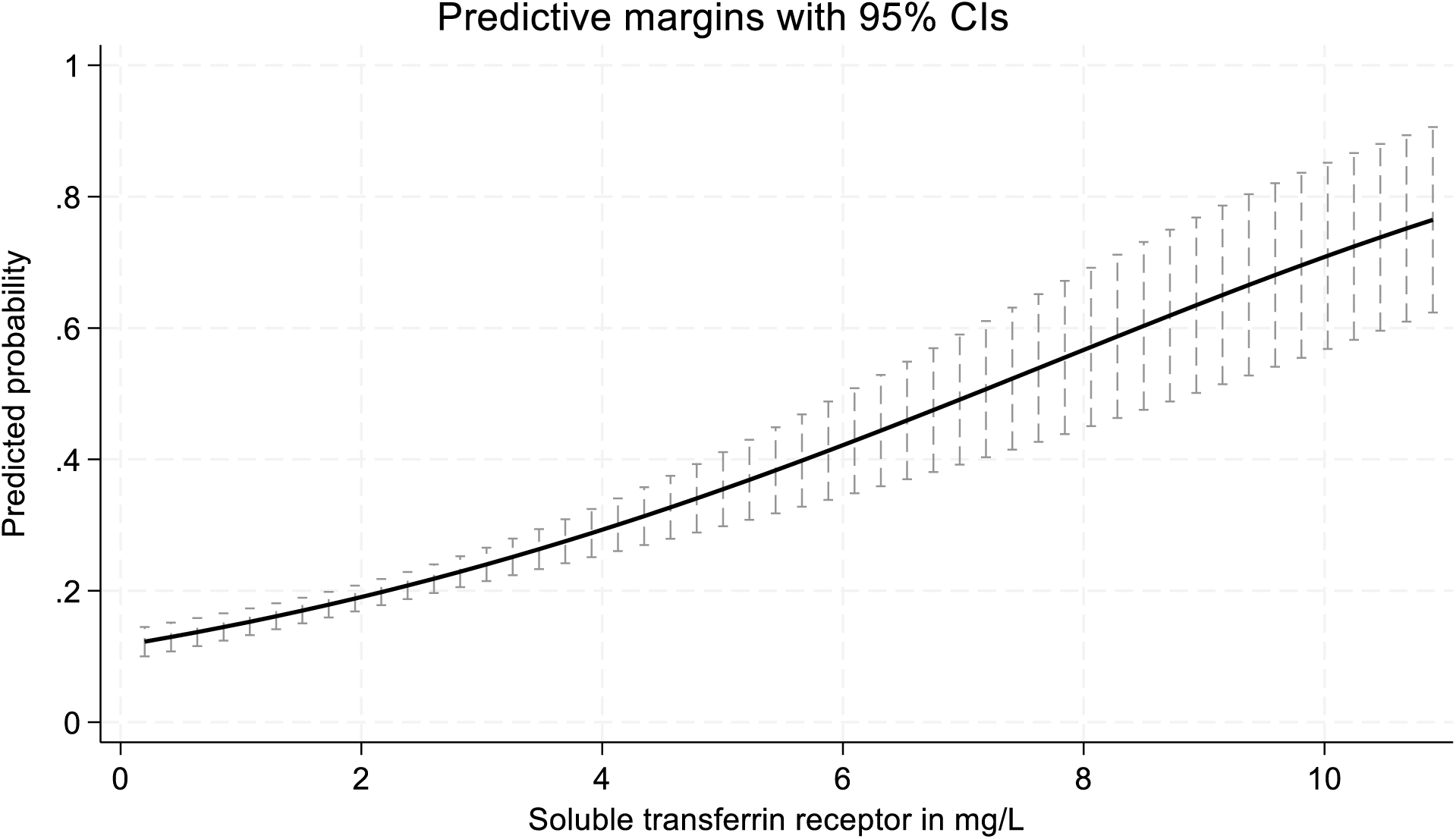
Linear relationship between AHF and sTfR

**Figure 4:**
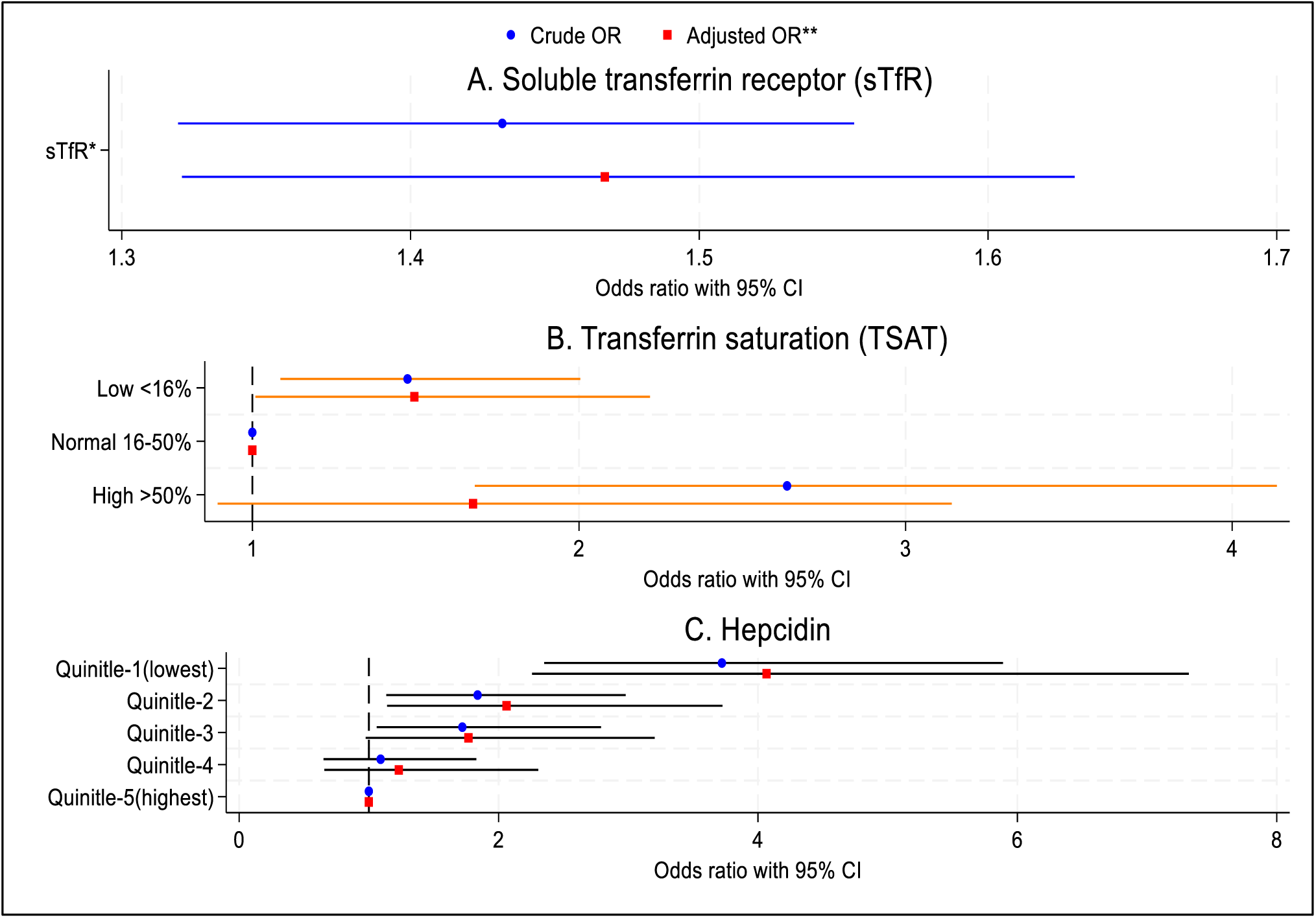
Association between AHF and maternal iron parameters Notes: different scales for the OR in the three figures. *OR in Figure A is odds of AHF per unit increase in the continuous variable, sTfR (no baseline/ reference). **Adjusted for woman’s age, BMI, religion, residence, below poverty line status, education level, tobacco, alcohol and betelnut consumption, smoking status, parity, antenatal care visits, HDP (not included in the hepcidin model, Figure C), medical comorbidities, multiple pregnancy, clustering by hospitals, days from conception to blood collection (TSAT model (Figure B) is additionally adjusted for blood transfusion in the antenatal period prior to the onset of AHF, and hepcidin (Figure C) model is additionally adjusted for serum CRP to control for inflammation).

Secondary analysis of ferritin using the ESC classification showed that the likelihood of AHF was significantly higher among women who had low ferritin levels compared with pregnant and postpartum women who had normal levels (see supplementary Table S6). Excluding 30 women with a known history of cardiac problem from the analyses did not change the findings (*results not shown*).

No significant interaction effects were observed between the iron parameters and HDP or medical comorbidities, except for hepcidin quintiles and HDP (p=0.0232). Similar to anemia, the multivariable models stratified by HDP, showed that the association between lower quintiles of hepcidin and AHF was only significant among women who did not have HDP, with the point estimates being much larger, but the odds of AHF did not vary significantly across the quintiles of hepcidin in women who had HDP (supplementary Table S7).

The association between the iron parameters and PPCM was similar to those observed for AHF including a positive linear association between PPCM and sTfR, higher odds of PPCM among women with low serum ferritin and low TSAT (see Table S8). However, the association between PPCM and quintiles of hepcidin was not significant, likely due to the smaller number of PPCM cases. Significant interaction effects were observed between sTfR and HDP. Analysis stratified by presence of HDP showed significant linear association between sTfR and PPCM in both groups and the effect sizes were also similar (supplementary Table S9).

### Effect of iron parameters on AHF with and without anemia

Stratified analysis by anemia showed independent associations between the iron parameters and both outcomes, AHF (Table-4) and PPCM (Table S10), suggestive of iron deficiency being associated with higher odds of the outcomes in women with and without anemia. The associations were similar to the findings of the main analysis. One observed difference was that among women who had no/mild anemia, the odds of AHF in the high TSAT group was around three-times the odds in the normal TSAT group even after adjusting for CRP and blood transfusion.

**Table 4:** Association of maternal iron parameters with AHF stratified by anemia.

| **Acute Heart Failure (AHF)** | **No/ mild anemia**<br>**(Hb ≥10gm/dL)** |  | **Moderate-severe anemia**<br>**(Hb <10 gm/dL)** |  |
| --- | --- | --- | --- | --- |
|  | **Adjusted OR**<br>**(95% CI)\*** | **p-value** | **Adjusted OR**<br>**(95% CI)\*** | **p-value** |
| **sTfR<sup>a</sup>** | 1.66 (1.27 – 2.16) | <0.001 | 1.30 (1.15 – 1.47) | <0.001 |
| **TSAT** |  |  |  |  |
| Low <16% | 1.28 (0.72 – 2.26) | 0.396 | 1.67 (0.88 – 3.19) | 0.118 |
| Normal 16-50% | 1 (ref) |  | 1 (ref) |  |
| High >50% | 3.04 (1.00 – 9.21) | 0.049 | 1.03 (0.43 – 2.47) | 0.946 |
| **Hepcidin quintiles** |  |  |  |  |
| Fifth (highest) | 1 (ref) |  | 1 (ref) |  |
| Fourth | 1.52 (0.66 – 3.52) | 0.325 | 1.21 (0.39 – 3.73) | 0.745 |
| Third | 2.12 (0.95 – 4.75) | 0.067 | 1.66 (0.58 – 4.69) | 0.343 |
| Second | 2.77 (1.23 – 6.24) | 0.014 | 1.51 (0.54 – 4.21) | 0.433 |
| First (lowest) | 4.00 (1.61 – 9.97) | 0.003 | 2.94 (1.07 – 8.06) | 0.036 |

<sup>a</sup>OR is odds of AHF per unit increase in the continuous variable (no baseline/ reference); \*Adjusted for woman's age, BMI, religion, residence, below poverty line status, education level, tobacco, alcohol and betelnut consumption, smoking status, parity, antenatal care visits, HDP (not included in the hepcidin model), medical comorbidities, multiple pregnancy, clustering by hospitals, days from conception to blood collection (TSAT model is additionally adjusted for blood transfusion in the antenatal period prior to the onset of AHF, and ferritin and hepcidin models are additionally adjusted for serum CRP to control for inflammation)

## Discussion

This multicenter case–control study from India provides the first robust evidence that iron deficiency and moderate-to-severe anemia significantly increase the risk of acute heart failure (AHF) in pregnant and postpartum women. Importantly, this risk was independent of hypertensive disorders of pregnancy (HDP) and pre-existing medical comorbidities including cardiac problems and remained consistent in the sub-group of women with peripartum cardiomyopathy, which is new onset heart failure with low LVEF. These findings identify iron deficiency as an important female-specific risk factor for pregnancy-associated AHF. Affected participants presented with severe symptoms and functional impairment (NYHA Class III / IV), and 30 women with AHF died during the study period. Echocardiographic findings revealed heterogeneous structural and functional abnormalities, consistent with the AHF syndrome, although pericardial effusion was present in one-third of cases. Of note, around 12% of postpartum women without suspected heart failure also showed abnormal echocardiographic features, and over one-third of them had iron deficiency and moderate-to-severe anemia. This suggests the possibility of subclinical cardiac dysfunction in this population. The absence of an association with abnormal hemoglobin variants further strengthens the inference that iron deficiency is the primary driver. Collectively, these findings support the role of iron deficiency and anemia as physiological stressors that impair normal cardiovascular adaptations in pregnancy and predispose to AHF.

Our results are consistent with previous observational studies from India^14–16^ and with our earlier systematic review^13^ but provide more comprehensive evidence by disentangling the effects of iron deficiency from anemia. Iron is essential for oxygen transport, DNA synthesis, cellular respiration, and electron transport, and it plays a particularly critical role in cardiac metabolism^32^. Outside the context of pregnancy, growing evidence indicates that iron deficiency independently impairs cardiac structure and function and may lead to cardiomyopathy^33,34^. A systematic review and meta-analysis of prospective studies showed an association between systemic iron deficiency and coronary artery disease and myocardial infarction in non-pregnant population^35^. A recent meta-analysis of six randomized controlled trials showed that treating iron deficiency in HF patients with intravenous iron significantly reduced cardiovascular events^36^. Experimental models suggest both causality, and point to mechanism involving myocardial iron depletion and ensuing metabolic and calcium signaling defects, all of which are reversible with iron repletion^37,38^.

To our knowledge, this large, multicenter study is the first worldwide to investigate in detail the association between AHF and iron deficiency in pregnancy and postpartum, using a comprehensive panel of iron biomarkers alongside echocardiographic evaluation. The research was needs-based driven by high case-fatality among pregnant and postpartum women in our study setting^10^, and guided by a-priori hypotheses from smaller studies^14–16^ and systematic review evidence^13^. While causality cannot be established in an observational design, a case–control approach was the most appropriate given the rarity of AHF, and the findings provide the strongest possible observational evidence to date. These results justify a future clinical trial of intravenous iron in pregnant and postpartum women with AHF, although such a trial would need to address ethical concerns about placebo use in iron-deficient women.

Controls were not matched to cases by gestational age; instead, all controls were recruited within 48 hours postpartum to reduce feasibility challenges and avoid misclassification. To minimize bias, both cases and controls were followed up through 6 weeks postpartum with standardized data collection across pregnancy. As explained, only a sub-group of the controls had echocardiography, due to limited clinical capacity at the start of the study. This does not constitute selection bias, as all controls recruited before October 2019 did not undergo evaluation. We included all controls, regardless of echocardiography status or abnormalities, which would bias the results toward the null rather than create spurious associations.

Although reverse causality cannot be fully excluded as iron deficiency is a comorbidity of heart failure^39^, the acute onset of HF in our cohort and the stronger association observed with anemia documented prior to AHF onset, mitigate this concern. Residual confounding is possible, although key confounders and effect modifiers such as sociodemographic factors, BMI, HDP and medical comorbidities were adjusted for. Blood parameters are influenced by time-period of pregnancy and postpartum, therefore we adjusted for the timing of blood collection in relation to women’s pregnancy and postpartum stage. Serum ferritin and hepcidin are influenced by inflammation which was accounted for by adjusting for CRP in the regression models. However, median CRP levels were 10 mg/L in both cases and controls, although the upper limit of the IQR was much higher in cases than controls. CRP physiologically increases after giving birth, especially in the first few days^40^; this could have biased the results towards null. Due to challenges in classifying iron deficiency using ferritin cut-offs, the association between ferritin and AHF was included as a secondary analysis, where we had to use different definitions for the exposure variable for cases and controls. While this is not recommended, using the same cut-off of <30 ng/mL would have led to significant misclassification of iron deficiency and overload in AHF cases (as shown in Table 3). Despite using the most recommended cut-offs for cases and controls we cannot completely exclude misclassification of low and high ferritin groups, but the results are consistent with those observed for sTfR, TSAT and hepcidin although apparently more extreme and therefore should be treated with caution. As serum ferritin is commonly used as a measure of iron in LMICs, there is need for research to identify appropriate cut-offs for this biomarker among pregnant women with heart failure.

Missing biomarker data was more frequent in cases due to critical illness and deaths. However, sensitivity analyses coding missing data as a separate category yielded results comparable to the primary analyses, suggesting robustness. Finally, BNP was not measured due to high cost, which may have led to under-detection of HF with preserved ejection fraction (HFpEF), although HFpEF in pregnancy is typically driven by HDP and afterload, and is usually accompanied by structural abnormalities captured on echocardiography^3^. Nevertheless, future studies should consider including measurements of BNP and exploring HFpEF in pregnant and postpartum women.

The heterogeneity of AHF pathophysiology underscores the need for comprehensive management strategies in pregnant and postpartum women. These should include screening for precipitating factors such as iron deficiency^12^, echocardiography to delineate cardiac dysfunction^12^, and management of the identified cardiac dysfunction and correction of iron deficiency with intravenous iron, which has been found to be safe in pregnancy^41^ and effective in patients with heart failure^36,39^. However, iron infusion should be targeted, as excess iron may cause cardiomyopathy. Stratified analyses by anemia suggested increased odds of AHF among non-anemia women with high ferritin and transferrin saturation, although residual confounding cannot be excluded. Additionally, long-term follow-up is essential to assess cardiac remodeling and to prevent future cardiovascular disease^3^.

Clinically, it is critical not to dismiss symptoms of AHF as normal pregnancy physiology or manifestations of anemia. Rapid recognition, referral, and initiation of treatment are essential, given the high case fatality in our study population. Delays in diagnosis, timely referral and treatment are known to be the major drivers in cardiac-related maternal death compared with noncardiac-related deaths^3^. It is important to identify sub-clinical heart failure or pre-heart failure based on recent recommendations^42^, which was likely present in around 12% of our controls. Telemedicine-based screening approaches, which we developed as part of this study^43^ may facilitate earlier identification of cardiac problems in pregnant women in low-resource settings.

At the policy level, national and international guidelines should expand beyond risk factors relevant to high-income countries to address conditions prevalent in LMICs. Iron deficiency is the most common nutritional deficiency in LMICs^44^. Iron deficiency anemia remains highly prevalent and represents a modifiable risk factor for maternal cardiac complications. Evidence indicates that up to 73% of adverse cardiac events in pregnancy are preventable^3^. Our findings call for greater equity in cardiovascular care for women in LMICs, with particular emphasis on screening and timely management of iron deficiency to reduce preventable maternal deaths.

## Acknowledgment

We gratefully acknowledge the commitment and dedication of the participants in this study.

## Author contributions

MN developed the concept and designed the study, led the overall work as chief investigator, analyzed the data, interpreted the findings, conducted the literature review and wrote the first draft of the paper. MN is both first and senior author. SSC is joint first author and contributed to the concept and design of the study, is master trainer for focused cardiac ultrasound used in the study, led the work in her institution as co-investigator, and edited the paper. AS contributed to analyzing the echocardiograms, provided expert advice as cardiologist in interpretation of the data, and edited the paper. VAI contributed to analyzing the echocardiograms, developing the study and editing the paper. SR, GD, CSV, AR, SDK, PM, FZ, IR are collaborators and investigators for the study, contributed to developing the study, and led the work in their respective institution. They also edited the paper. RD is the MaatHRI program manager, supervised study staff and data collection and edited the paper. CO supported the statistical analysis section and edited the paper. MA and SK contributed to analyzing the echocardiograms and edited the paper. PL contributed to designing the study and edited the paper. SLL advised on the iron homeostasis aspects of the study and edited the paper. BC contributed to designing the study and editing the paper. JA contributed to designing the study, interpreting the results and editing the paper.

